# Novel Alzheimer’s disease genes and epistasis identified using machine learning GWAS platform

**DOI:** 10.1101/2023.10.04.23296569

**Authors:** Mischa Lundberg, Letitia M.F. Sng, Piotr Szul, Rob Dunne, Arash Bayat, Samantha C. Burnham, Denis C. Bauer, Natalie A. Twine, the Alzheimer’s Disease Neuroimaging Initiative

## Abstract

Alzheimer’s disease (AD) is a complex genetic disease, and variants identified through genome-wide association studies (GWAS) explain only part of its heritability. Epistasis has been proposed as a major contributor to this ‘missing heritability’, however, many current methods are limited to only modelling additive effects. We use VariantSpark, a machine learning (ML) approach to GWAS, and BitEpi, a tool for epistasis detection, to identify AD associated variants and interactions across two independent cohorts, ADNI and UK Biobank. By incorporating significant epistatic interactions, we captured 10.41% more phenotypic variance than logistic regression (LR). We validate the well-established AD loci, *APOE*, and identify two novel genome-wide significant AD associated loci in both cohorts, *SH3BP4* and *SASH1*, which are also in significant epistatic interactions with *APOE*. We show that the *SH3BP4* SNP has a modulating effect on the known pathogenic *APOE* SNP, demonstrating a possible protective mechanism against AD. *SASH1* is involved in a triplet interaction with pathogenic *APOE* SNP and *ACOT11,* where the *SASH1* SNP lowered the pathogenic interaction effect between *ACOT11* and *APOE*. Finally, we demonstrate that VariantSpark detects disease associations with 80% fewer controls than LR, unlocking discoveries in well annotated but smaller cohorts.

## Introduction

Alzheimer’s disease (AD) is the most common form of dementia and predominantly affects individuals over 65 [1]. The vast majority (99%) of AD cases are late onset (LOAD) and are driven by multiple genetic and environmental influences, with genetics accounting for between 53% and 80% of total phenotypic variance [2–4]. The heritability of LOAD is predominantly carried by the *APOE* locus, which explains about 25% of the total heritability of the disease [5]. In addition to *APOE*, large-scale genome-wide association study (GWAS) meta-analyses identified 40 [6] and 75 additional risk loci [7], but more than 30% of genetic variability remains unknown [3]. Recent studies [8,9] predict that there are 100-1,000 causal variants with modest effects associated with LOAD, of which only a small proportion have been identified.

Part of the missing heritability in LOAD might be explained by non-additive interactions [10], which are ignored by GWAS studies. Indeed, a genome-wide replicated scan has found epistasis to be a ubiquitous phenomenon across multiple phenotypes [11]. Epistatic interactions have long been implicated in complex genetic disease, including neurological diseases [12] and LOAD itself [13]. However, due to the computational complexity of finding genome-wide gene-gene interactions, the search were limited to candidate gene approaches [13–17], or genome-wide approaches exploring interactions between *APOE* and other risk loci [18].

Using the ML platform VariantSpark [19], we overcome the shortcomings of traditional statistical GWAS approaches and computationally limited epistatis discovery tools and identify genome-wide variants associated with LOAD and AD in both the Alzheimer’s Disease Neuroimaging Initiative (ADNI) cohort (512 cases, 272 controls) [20] and the UK Biobank (UKBB) cohort (704 cases, up to 6869 controls) [21]. Using a novel false discovery rate (FDR) method [22], we are able to use VariantSpark’s random-forest-based feature selection approach to narrow down the genome-wide search space to the subset of variants enriched with epistatic interactions. We then apply BitEpi [23] to perform an exhaustive search of this subset to annotate pairwise and higher-order, statistically significant interactions between the variants. We also explore the proportion of phenotypic variance captured by VariantSpark versus the traditional logistic regression (LR) methods. Finally, we demonstrate that VariantSpark has improved sensitivity to detect signal with fewer control samples compared with LR approaches.

## Results

### A. VariantSpark identifies known AD loci across two independent cohorts

Using the ML genomics platform VariantSpark [19], and a novel RFlocalfdr approach [22], we identified genetic variants that are both marginally and interactively associated in two independent AD cohorts, UKBB and ADNI (7,573 and 784 samples of 4.5M SNPs each). Because of these two types of associations, we expect to find more significant variants than a LR approach at <5% FDR.

We identified 104 SNPs (53 independent) to be significantly associated with AD in the UKBB cohort (Table 1, Figure 1, Supplementary Table S1) and 207 significantly associated SNPs (124 independent) in the ADNI cohort (Figure 1, Supplementary Table S2). When we compared these associations with those associated with AD in the GWAS Catalog (trait ID ‘MONDO_0004975, accessed 16/05/22)[24] using locus bins, we observed a 70% overlap with the significant SNPs identified in both the UKBB (72/104) and ADNI cohort (145/207), with 31 out of the 53 independent UKBB SNPs (58.49%) and 82/124 (66.13%) of the independent ADNI SNPs (Table 1 and Supplementary Table 2).

**Figure 1.**
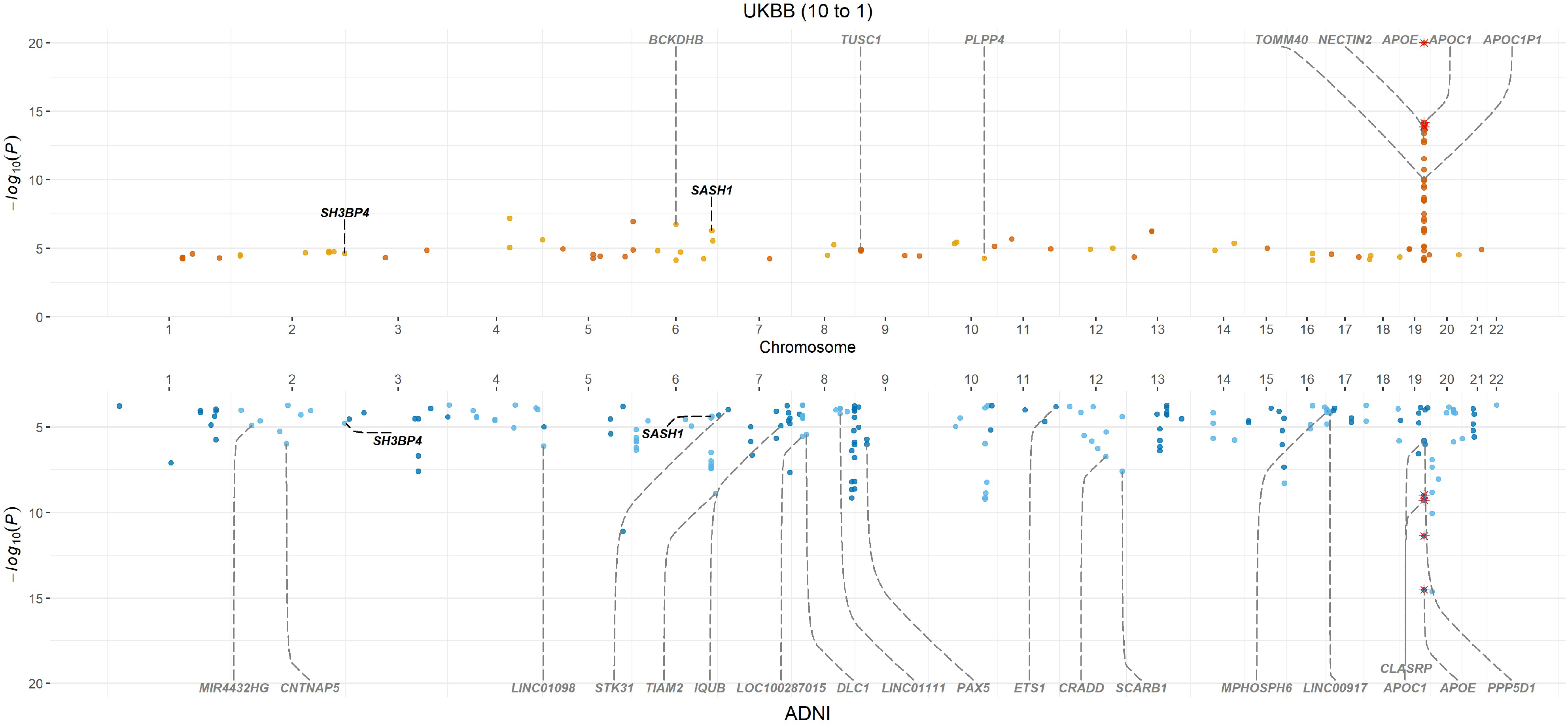
Miami plot showing significant SNPs identified by VariantSpark in UK Biobank (10 controls to 1 case) (top) and ADNI (bottom) cohorts. Red asterisks mark those variants that have been replicated by position (only independent variants) between the two cohorts. Annotation (black) represents gene annotations that are novel and replicated between the two cohorts. Annotations in grey represent previously identified variants.

**Table 1.** Annotated statistically significant independent SNPs identified using VariantSpark and the UKBB cohort. Replication status to ADNI validation cohort and GWAS Catalog included.

| Chr | Pos | Alt | RSID | P-value | Gene | ADNI | GWAS Catalog |
| --- | --- | --- | --- | --- | --- | --- | --- |
| 1 | 174049377 | C | rs72711440 | 4.36E+09 | RABGAP1L-DT | FALSE | FALSE |
| 1 | 193137934 | C | rs139963893 | 2.50E+09 | CDC73 | FALSE | TRUE |
| 1 | 244007787 | A | rs75965920 | 5.16E+09 | AKT3 | FALSE | FALSE |
| 2 | 38520113 | A | rs56302953 | 3.05E+09 | ATL2 | FALSE | TRUE |
| 2 | 205966824 | G | rs116192932 | 1.64E+09 | PARD3B | FALSE | TRUE |
| 2 | 215673948 | C | rs71579843 | 1.70E+09 | BARD1 | FALSE | FALSE |
| 2 | 235751287 | G | rs114656810 | 2.29E+09 | SH3BP4 | TRUE | FALSE |
| 3 | 76534878 | C | rs74801337 | 4.85E+09 | ROBO2 | FALSE | FALSE |
| 3 | 154755590 | C | rs9829241 | 1.39E+09 | MME | FALSE | FALSE |
| 4 | 116269757 | A | rs147896092 | 6.36E+05 | NDST4 | FALSE | FALSE |
| 4 | 178201067 | G | rs76175875 | 2.34E+08 | NEIL3 | FALSE | TRUE |
| 5 | 36648950 | C | rs115172522 | 1.07E+09 | SLC1A3 | FALSE | TRUE |
| 5 | 93761812 | G | rs112217406 | 2.73E+09 | KIAA0825 | FALSE | FALSE |
| 5 | 153560523 | G | rs71585927 | 3.90E+09 | GALNT10 | FALSE | TRUE |
| 5 | 168782459 | C | rs78269616 | 1.07E+07 | SLIT3 | FALSE | FALSE |
| 6 | 46454250 | G | rs114866534 | 1.49E+09 | RCAN2 | FALSE | TRUE |
| 6 | 81120137 | G | rs117103821 | 1.75E+07 | BCKDHB | FALSE | TRUE |
| 6 | 133182939 | NA | rs57823471 | 5.59E+09 | NA | TRUE | TRUE |
| 6 | 148512131 | C | rs117160741 | 5.15E+07 | SASH1 | TRUE | TRUE |
| 6 | 150085133 | A | rs117243801 | 2.84E+08 | PCMT1 | FALSE | TRUE |
| 7 | 101930310 | C | rs11552019 | 5.76E+09 | SH2B2 | FALSE | TRUE |
| 8 | 52718273 | T | rs112585504 | 3.10E+09 | PXDNL | FALSE | FALSE |
| 8 | 65042594 | T | rs78789176 | 5.43E+08 | LINC01414 | FALSE | TRUE |
| 9 | 25640336 | A | rs77057081 | 1.16E+09 | TUSC1 | FALSE | TRUE |
| 9 | 108857477 | C | rs117992330 | 3.40E+09 | TMEM38B | FALSE | TRUE |
| 9 | 136144593 | G | rs66697526 | 3.65E+08 | ABO | FALSE | TRUE |
| 10 | 66463703 | C | rs189699806 | 4.56E+07 | ANXA2P3 | FALSE | FALSE |
| 10 | 70366297 | C | rs61868095 | 3.59E+08 | TET1 | FALSE | TRUE |
| 10 | 122310126 | C | rs79486209 | 5.24E+09 | PLPP4 | FALSE | FALSE |
| 11 | 12566857 | C | rs76154502 | 7.12E+08 | PARVA | FALSE | TRUE |
| 11 | 44909137 | C | rs78150932 | 2.09E+07 | TSPAN18 | FALSE | TRUE |
| 11 | 118613605 | A | rs139648410 | 1.08E+09 | DDX6 | FALSE | FALSE |
| 12 | 65040623 | G | rs79774071 | 1.12E+09 | RASSF3 | FALSE | TRUE |
| 12 | 107592329 | G | rs75378184 | 9.57E+08 | CRY1 | FALSE | TRUE |
| 13 | 22104766 | T | rs116942699 | 4.31E+09 | MICU2 | FALSE | FALSE |
| 13 | 55121441 | G | rs117658626 | 5.37E+07 | MIR1297 | FALSE | FALSE |
| 14 | 62969759 | G | rs79473324 | 1.39E+09 | KCNH5 | FALSE | FALSE |
| 14 | 99261321 | C | rs142983586 | 4.35E+07 | C14orf177 | FALSE | TRUE |
| 16 | 53388460 | A | rs2908783 | 2.29E+09 | LOC643802 | FALSE | FALSE |
| 17 | 2761430 | C | rs74252350 | 2.63E+09 | RAP1GAP2 | FALSE | FALSE |
| 17 | 54490063 | C | rs147441895 | 4.16E+08 | ANKFN1 | FALSE | FALSE |
| 18 | 22582278 | T | rs79110874 | 3.25E+09 | LINC01894 | FALSE | FALSE |
| 18 | 77685936 | G | rs113702893 | 4.25E+09 | SLC66A2 | FALSE | FALSE |
| 19 | 17262529 | C | rs117951200 | 1.12E+09 | MYO9B | FALSE | TRUE |
| 19 | 45411941 | T | rs429358 | 0.00E+00 | APOE | TRUE | TRUE |
| 19 | 55511927 | G | rs1560714 | 2.98E+09 | NLRP2 | FALSE | TRUE |
| 21 | 36714721 | A | rs2834914 | 1.22E+09 | LOC100506403 | FALSE | FALSE |

As expected, the *APOE* loci was identified in both ADNI and UKBB cohorts (Supplementary Tables 1 and 2). To evaluate the functional context of the other significantly associated independent variants, we performed functional enrichment analysis using MAGMA. Gene-set analysis (Supplementary Table S4) identified 9 (ADNI) and 3 (UKBB) gene sets significantly associated (after Bonferroni correction). Many of the significant gene sets and those with suggestive significance levels (P < 0.05) fell into the categories of transmembrane and metal ion transport proteins (known to be key in neuronal signalling in the brain). Tissue expression analysis using MAGMA and GTEX (Supplementary Table S5, Supplementary Table S6) revealed brain tissues to be the most highly ranked, although they did not pass Bonferroni correction.

### B. VariantSpark identifies novel loci associated with AD

We next investigated which loci replicated between the two independent cohorts. Despite the phenotypic heterogeneity across the two cohorts, we replicated three independent, significantly associated genes, *APOE* (rs429358), *SASH1*, and *SH3BP4* (Table 1 and Supplementary Table S2). It is important to note that the significance threshold for the RFlocalfdr is 0.05 compared to the traditional genome-wide significance threshold of *P* < 5×10^-8^, which needs to correct for multiple tests. Both thresholds, RFlocalfdr for VariantSpark and *P* < 5×10^-8^ for logistic regression, control for Type 1 error and correct for the multiple testing burden. For further information, see Methods section.

Both *SASH1* and *SH3BP4* were novel to our study and were not yet present in the GWAS Catalog SNPs, although there is a marginally associated SNP (rs9390537, χ^2^-*p* = 8.17×10^-6^) mapping to an intergenic region 91,233 bp upstream of *SASH1* associated to AD [25] and another marginally associated SNP (rs66501349, χ^2^-*p* = 2×10^-6^) intergenic to *SH3BP4* and *CEP19P1* associated to poorer cognitive function [26]. The corresponding rsIDs from the UKBB cohort are rs117160741 (Chr 6:148512131) for *SASH1* and rs114656810 (Chr 2:235751287) for *SH3BP4* (Supplementary Table S1). Both are intergenic and located upstream of the genes. Similarly, the rsIDs from the ADNI cohort are rs9918382 (Chr 6:148265029), an intergenic variant located upstream of *SASH1,* while rs6711272 (Chr 2:235131361 is an intergenic variant located downstream of *SH3BP4* (Supplementary Table S2).

*SASH1* (SAM and SH3 domain-containing 1) encodes a scaffold protein, which is ubiquitously expressed, including in brain tissues and is also a positive regulator of the NF-_k_B signalling pathway through the activation of *TLR4* [27]. *SH3BP4* (SH3 domain binding protein 4) encodes a protein involved in the amino acid-induced TOR signalling pathway [28]. Both *SASH1* and *SH3BP4* are membrane bound phosphoproteins with SH3 domains.

### C. BitEpi identifies novel interactions between known and novel AD genes

BitEpi was used to identify epistatic interactions between significantly associated variants in both cohorts. The β and α metrics, reflecting association power and interaction effect respectively, were used to select interactions that were strongly associated to the AD phenotype due to an epistatic effect. We identified 37 interactions with significant β and α values in the UKBB cohort, of which 17 were 2-SNP, 16 were 3-SNP, and 4 were 4-SNP interactions (Figure 2, Supplementary Table S7). Using the ADNI cohort, we identified 58 interactions with significant β and α values, 39 were 2-SNP, 17 were 3-SNP and 2 were 4-SNP interactions (Figure 3, Supplementary Table S8). Interestingly, the two replicating AD associated genes, *SASH1* and *SH3BP4*, were involved in epistatic interactions.

**Figure 2.**
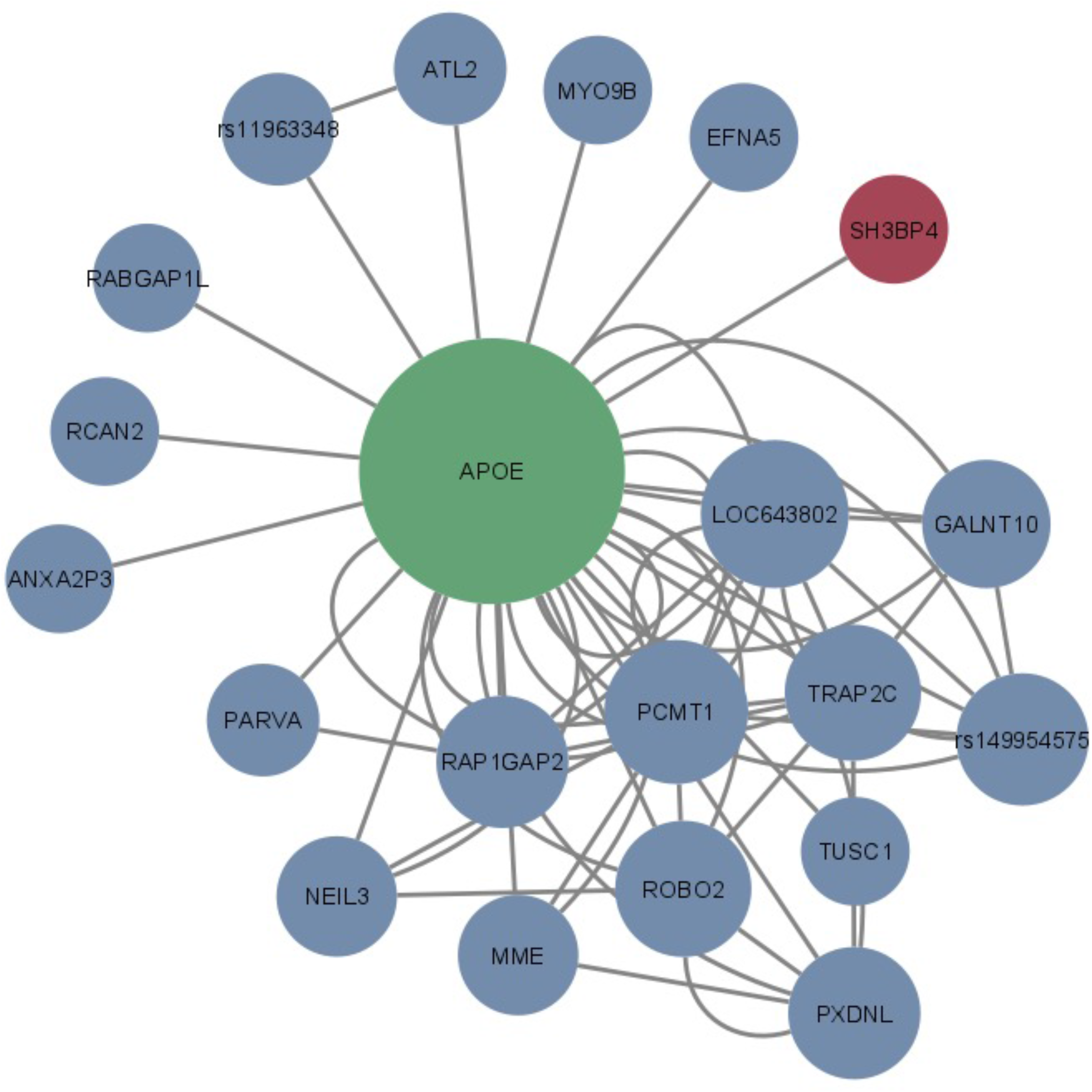
Network diagram of significant BitEpi interactions from UK Biobank cohort. Nodes in green are known AD associations, in red is the novel gene replicated in this study, and in blue are variants which are novel but unreplicated. All 2-SNP, 3-SNP, and 4-SNP interactions are included. Size of nodes are representative of node degree calculated from the NetworkAnalyzer plug-in in Cytoscape.

**Figure 3.**
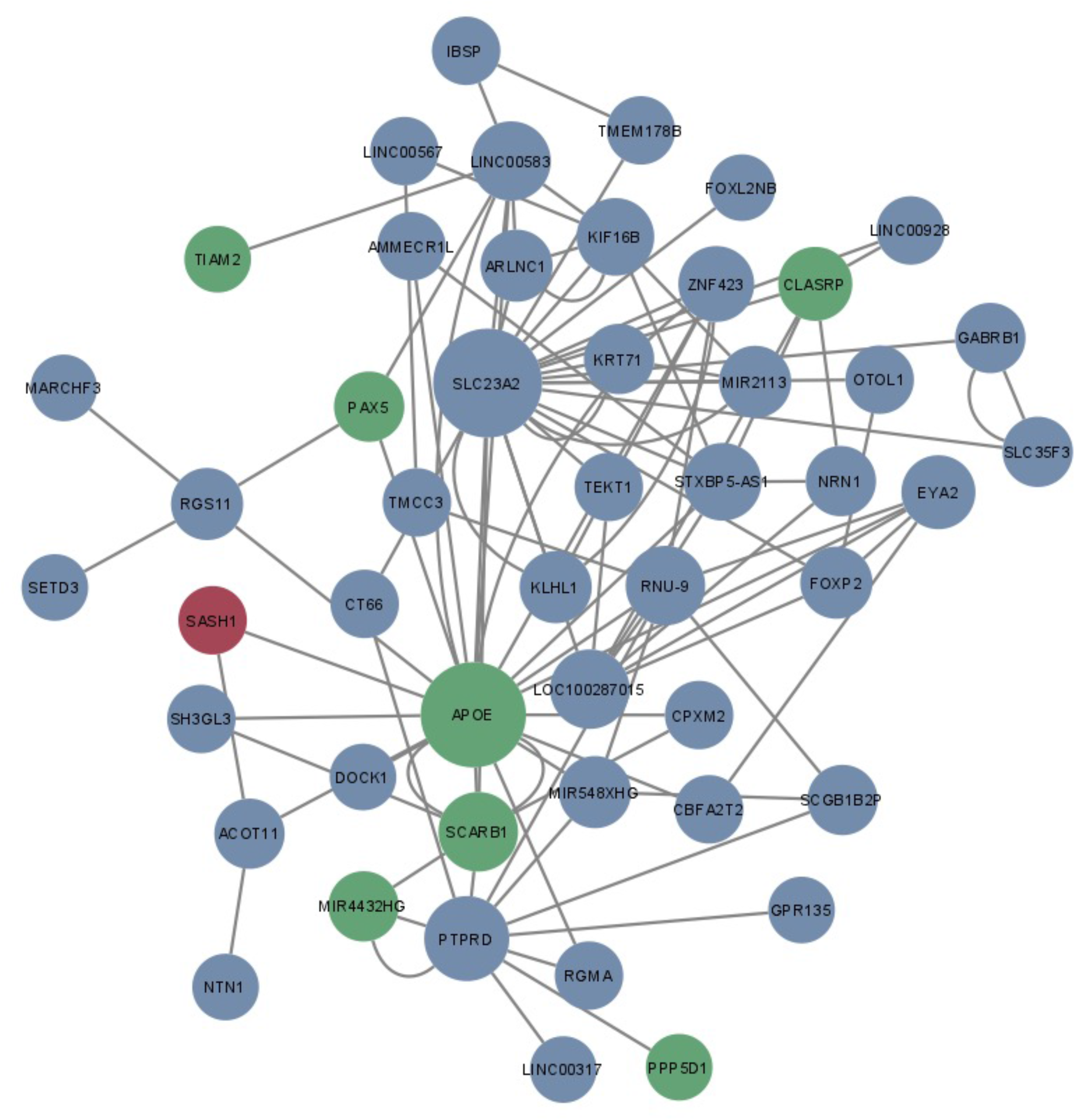
Network diagram of significant BitEpi interactions from ADNI cohort. Nodes in green are known AD associations, in red is the novel gene replicated in this study, and in blue are variants which are novel but unreplicated. All 2-SNP, 3-SNP, and 4-SNP interactions are included. Size of nodes are representative of node degree calculated from the NetworkAnalyzer plug-in in Cytoscape.

In the UKBB cohort, the SNP (rs114656810) mapping to *SH3BP4* was found to interact with rs429358, which is a reported pathogenic *APOE* SNP in ClinVar [29], where the alternate ‘C’ allele plays a part in the high AD-risk APOE-ε4 isoform. This pairwise interaction was interrogated to identify the genotype combinations associated with AD (Supplementary Table S9). Due to the low number of samples with the homozygous alternate genotype (AA) of *SH3BP4* SNP, we reduced the genotypes to two classes; presence or absence of the alternate ‘A’ allele. In the absence of the alternate *SH3BP4* SNP allele, there was no absolute difference in control rates between the *SH3BP4*x*APOE* interaction and the *APOE* SNP alone (Figure 4A). This indicates a limited effect of the homozygous reference genotype of rs114656810 on AD. However, with the presence of the alternate allele of the *SH3BP4* SNP, the pathogenic effect of the *APOE* C allele is modulated (Figure 4A), suggesting that *SH3BP4* may have a protective mechanism against AD for carriers of the *APOE* ‘CC’ genotype. In the ADNI cohort, this pairwise interaction between *SH3BP4* and *APOE* was marginally significant but did not pass Bonferroni correction.

**Figure 4.**
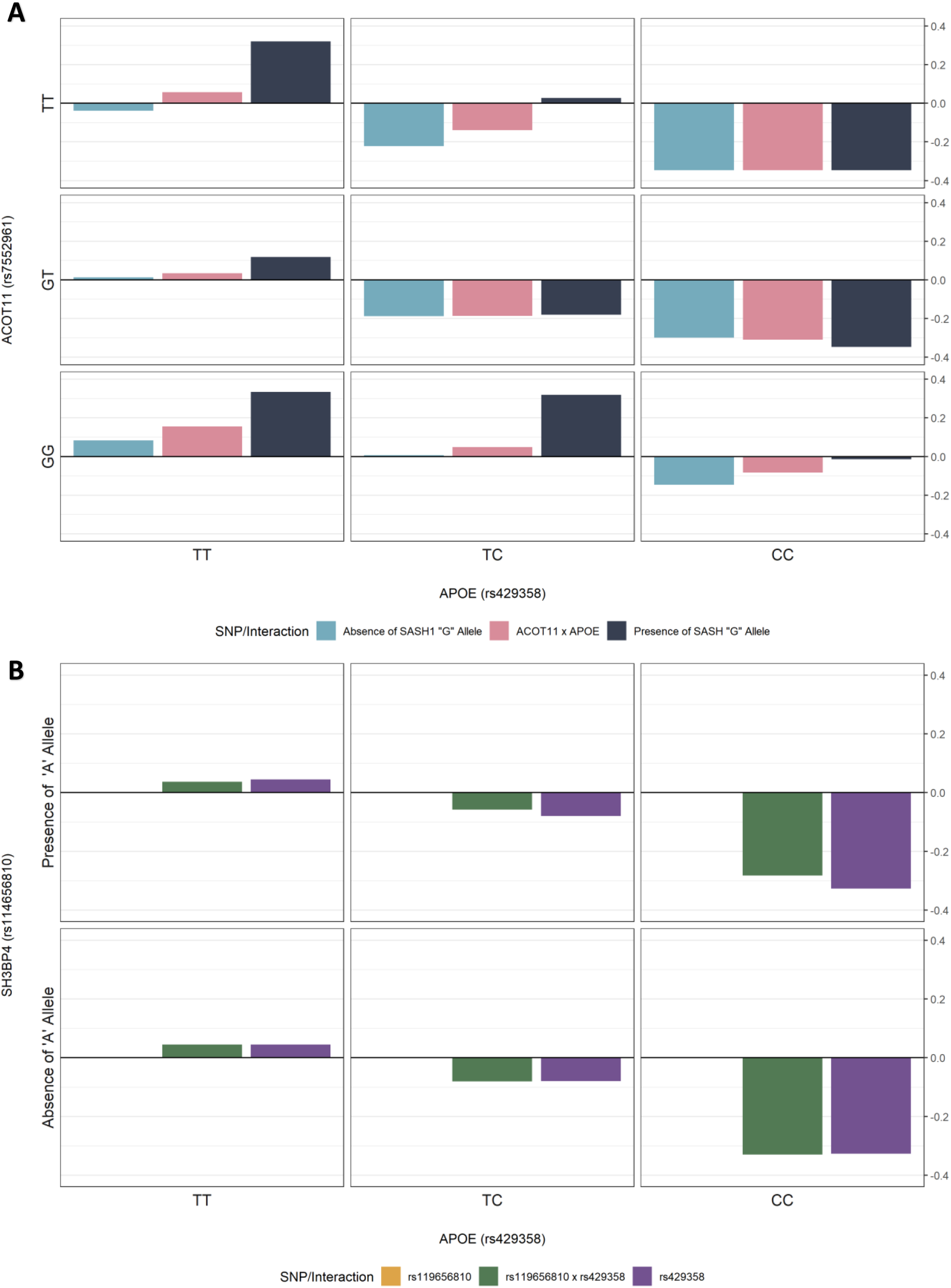
Relative control rates of the interactions **(A)** rs119656810 (*SH3BP4*) and rs429358 (*APOE*) in the UK Biobank cohort, and **(B)** rs7552961 (*ACOT11*), rs9918382 (*SASH1*), and rs429358 (*APOE*) in the ADNI cohort. Relative control rates were calculated as the difference between control rates of each genotype combination and the control rate of the entire cohort. Due to sample size restrictions, the rs119656810 SNP and the rs9918382 SNP was reduced to two categories; presence or absence of its alternate allele. There is evidence of a modulating effect of the alternate allele of rs119656810 on the *APOE-e4* (rs429358 CC) genotype as seen from the increase in relative control rates in the top middle and top right cells in **(A)**. There is evidence of a protective effect of alternate allele of rs9918382 on the *ACOT11* x *APOE* genotypes as seen from the increase in relative control rates in the top middle cell and the bottom right cell in **(B)**. However, there is no evidence of the same effect for the *APOE-e4* (rs429358 CC) genotype in an interaction with the *ACOT11* alternate allele (rs7552961).

In the ADNI cohort, the SNP rs9918382 mapping to *SASH1* was involved in a triplet interaction with the same pathogenic APOE SNP, rs429358. The other SNP, rs7552961, in the triplet maps to *ACOT11*, has been shown to be associated to mild cognitive decline [30]. This triplet interaction was also examined further (Supplementary Table S10). Again, due to the low numbers of samples with the homozygous alternate genotype of rs9918382 (*n* = 15), the genotype was reduced to two classes; presence or absence of the alternate ‘G’ allele. Figure 4B shows that the alternate ‘G’ allele of the *SASH1* SNP has a protective effect, reversing the pathogenic interaction effect of the rs7552961 (*ACOT11*) TT genotype and rs429358 (*APOE*) TC genotype increasing the relative control rate from –0.139 to 0.028 (Supplementary Table S10). However, when the alternate *ACOT11* allele (G) is present with the *APOE* CC genotype, the *SASH1* SNP has no effect. In fact, none of the possible pairwise interactions between these three genotypes passed significance for the α metric, which suggests that the association to AD was carried by the interaction of all three SNPs. This highlights the complexity and difficulty of detecting epistatic interactions, where exacerbating or protective properties are exerted through specific combinations of genotypes.

### E. VariantSpark can detect more disease associated signal than logistic regression

Next, we compared VariantSpark with the more traditional GWAS approach implemented in PLINK’s logistic regression (LR) to estimate the power to detect disease associated signal with limited control samples. To do this, in addition to using the ADNI cohort, we subset two datasets from the UKBB cohort: the first contained a ratio of 10 controls to 1 case (UKBB10to1) and the second with 2 controls to 1 case (UKBB2to1).

Using LR, we did identify multiple variants at suggestive significance levels using the ADNI cohort (ranging from χ^2^-*p* = 8.34×10^-8^ to χ^2^-*p* = 2.63×10^-6^), all falling into the *APOE* locus (Chr19:45,326,217 to Chr19:45,445,517). Based on the UKBB cohort, we identified three significantly independent associated SNPs in UKBB10to1 (127 in total) (Supplementary Table S3) and one significantly independent associated SNP in UKBB2to1 (74 in total) (Supplementary Table S11). All SNPs found using LR fell within the *APOE* locus (Chr19:45,326,217 to Chr19:45,445,517).

In contrast, VariantSpark identified associations outside of the *APOE* region such as rs79486209 on chromosome 10 which mapped to *PLPP4*, a gene previously associated with AD [31]. VariantSpark identified 53 significantly associated independent SNPs (104 in total) in UKBB10to1 (Table 1) and 20 significantly associated independent SNPs (69 in total) in UKBB2to1 (Supplementary Table S12).

This demonstrates we have 15% (1/3 vs. 20/53) more power to detect disease associated variants with 80% fewer (2 vs. 10) controls using VariantSpark compared with a LR approach.

### F. VariantSpark captures more phenotypic variance in AD than Logistic Regression

A key goal of this study was to explore whether epistasis can explain some of the missing heritability that is well documented in AD [2–4]. To this end, we measured the proportion of phenotypic variance captured by genetic variants identified in the UKBB cohort using Nagelkerke’s pseudo-R^2^ and fitting three LR models with: Firstly, significant and independent SNPs identified by LR (n = 3). Secondly, significant and independent SNPs identified by VariantSpark (n = 53). Thirdly, significant and independent SNPs identified by VariantSpark with significant interactions identified by BitEpi (n = 122).

Within the UKBB cohort, the VariantSpark-BitEpi model (model (3)) captured the highest variance explained at 23.18% compared to model (2) without the BitEpi interactions at 17.12% and model (1) the LR SNPs at 12.77% (Supplementary Fig. S2). To test whether the performance increase of the VariantSpark-BitEpi model was driven by its additional variables, we calculated an empirical *P*-value. We fitted 1,000 models containing the 3 LR SNPs as well as 50 randomly selected SNPs and 69 interactions to emulate the degrees of freedom of the VariantSpark-BitEpi model (3). As shown in Supplemental Fig. S2, these models achieved an average pseudo-R^2^ of 19.33%, outperforming the models with fewer predictors (models (1) and (2)). In contrast, VariantSpark-BitEpi’s model had a small but significant (*p* = 0.006) performance improvement over the random models (23.18% vs 19.33%), confirming that additional signal was captured. We make a similar observation for these models when tested on the independent ADNI cohort. LR (model 1) captured 7.09% while the random models captured 25% on average and VariantSpark-BitEpi (model 3) achieved 27.20%. The increase in variance explained on the ADNI set is likely due to an easier signal, which is predominantly driven by *APOE* (as observed in Section C).

These findings indicate that VariantSpark-identified SNPs and BitEpi-identified epistatic interactions together explain up to 10.41% more phenotypic variance in AD than traditional LR approaches that focus only on marginal effects. This also aligns with previous studies where the addition of 87 marginal effect SNPs (without *APOE*) explained only 2.1% more variance [32] and 2,042,105 SNPs (without known AD SNPs) accounted for 25.3% variance [3]. Taken together, these results suggest that epistatic interactions across the genome play a part in AD aetiology and should be accounted for when developing therapeutics and genetic risk scores.

### G. Transcriptome-wide association (TWAS) lookup of SASH1 and SH3BP4

Finally, we looked at transcriptomic level information of the mapped genes *SASH1* and *SH3BP4* as previous studies [33,34] have shown that this can add confidence that GWAS-identified genes are capturing actual disease-related signal. Using the TWAS-hub [35], *SASH1* showed strong evidence (ENET-*P* = 7.5×10^-9^) of involvement in the prefrontal cortex tissue and a strong association with “Alzheimer’s Disease (in father)” (Supplementary Table S13). In contrast, *SH3BP4* showed an association with nerve tibial tissue at non-suggestive levels for Alzheimer’s Disease (Supplementary Table S14). Another resource used were the gene expression tests built into FUMA [36] using GTEx v8 [37] data. In this analysis, both *SASH1* and *SH3BP4* showed increased expression levels in brain tissue (Supplementary Fig S3).

## Discussion

Using VariantSpark, a ML approach to GWAS, we have identified two novel genes, *SASH1* and *SH3BP4,* to be associated with AD reaching genome-wide significance..

*SASH1* is a known tumour suppressor protein that has been shown to be differentially expressed between AD and control samples [38,39]. Furthermore, a previous study found SNP rs9390537 (located 91,233bp upstream of *SASH1)* to be nominally associated to LOAD (χ^2^-*p* = 8.17×10^-6^) [25]. Indeed, it is a nominated AD drug target in the Agora database, a database curated by AD researchers from the accelerating medicine partnership-Alzheimer’s disease consortium and other research teams.

*SH3BP4* or transferrin trafficking protein (*TTP*) interacts with endocytic proteins including clathrin, dynamin, and the transferrin receptor [40] and is involved in the aminal acid-Rag GTPase-mTORC1 signalling pathway. It is a central link between Akt signalling and cell-matrix adhesion regulation [28]. Although *SH3BP4* has no established link to AD, a SNP (rs66501349, intergenic to *SH3BP4* and *CEP19P1*) has been marginally associated to poorer cognitive function (χ^2^-*p* = 2×10^-6^) [26] and its interactor dynamin has strong evidence of a role in AD pathophysiology [41,42]. In particular, the expression of gene *DNM2* was significantly decreased in AD patients, and neuronal cell lines transfected with dominant negative *DNM* genes were observed to have an accumulation of APP and increased Aβ secretion [43].

The key contribution of our work is adding the lens of epistasis to association. We identified a total of 95 epistatic interactions, including 2-SNP, 3-SNP and 4-SNP interactions associated with AD, in two independent cohorts. This elevated the previously only nominally associated *SASH1* [25] to pass FDR significance when its interaction with *ACOT11* and *APOE* is accounted for. Specifically, our epistasis analysis revealed that the alternate ‘G’ allele of *SASH1* SNP rs9918382 appears to have a protective effect against AD as it reverses the pathogenic effect of *ACTO11* rs7552961 ‘TT’ and *APOE* rs429358 ‘TC’ genotype combination (Supplementary Fig. S3). However, this modulating effect was not found in the presence of two copies of the pathogenic *APOE* ‘C’ allele (rs429358, Supplementary Fig. S3). This result is consistent with co-expression patterns found between AD and control brains [44] and the high expression levels of *SASH1* in pre-frontal cortex tissue in the TWAS-hub. Taken together, it is likely that *SASH1* plays a role in AD pathophysiology and warrants further investigations.

Although, most of our identified epistasis is concentrated between *APOE* and a small number of other loci, our methodology can explore genome-wide epistasis in an unbiased manner, unlike previous studies [45,46]. Additionally, a genome-wide search allows for the identification of epistasis in non-coding regions of the genome which have empirically demonstrated to effect gene expression [47].

For example, our epistasis analysis revealed a modulating effect of the alternate allele of SNP *rs*119656810 (*SH3BP4*) on the *APOE* locus. A possible explanation for this effect is that *SH3BP4*’s ability to regulate the activity of dynamin [40], it enables the processing of amyloid β protein precursors resulting in lower levels of Aβ depositions and AD pathology. Together, *SH3BP4* is a novel gene that may play a role in AD pathophysiology through its pathway mechanisms and in combination with *APOE*.

While VariantSpark identified *SH3BP4* and *SASH1* in both cohorts due to their cumulative additive and epistatic effects on AD, the exact epistatic interactions they are involved in were not replicated although, *SH3BP4*-*APOE* showed marginal significance. This is likely due to the varying number of individuals who might have this exact modulating disease physiology and genotype combinations across the two cohorts. This illustrates the benefits of using VariantSpark instead of traditional LR models on binary traits with potential polygenic interactions, like Alzheimer’s disease.

Using VariantSpark, we were also able to detect disease genes with fewer controls than traditional approaches. This is relevant as a recent study calculates 10,000,000 cases would be needed for a traditional GWAS to find significant SNPs explaining 50% of Alzheimer’s disease heritability [48]. Even for large initiatives such as FinnGen or 23andMe, such numbers are hard to achieve. Our method offers an alternative and enables discoveries in smaller but well annotated cohorts for AD and other genetic studies.

The limitations to our study are as follows: Firstly, ADNI used whole genome sequencing mapped to the GRCh38 reference genome, while the UKBB used array technology mapped to the GRCh37 reference genome resulting in the final set of 4.5 million common SNPs which was around 50% of the total number of SNPs for both cohorts. Secondly, the ADNI and UKBB cohorts are both different ascertainments. Particularly, UKBB is a relatively healthy volunteer cohort and contained a mix of AD phenotypes while ADNI recruited patients based on their health status and included samples with mild cognitive impairment to maximise sample size but is only an AD-proxy phenotype. Lastly, the ADNI cohort was substantially smaller than the UKBB cohort, with 784 samples, compared to 7582 samples. These three factors in combination limit our power to discover and replicate disease variants and epistatic interactions across cohorts. Furthermore, we restricted our samples in both cohorts to those of European descent as is commonplace [49]. However, it has been shown that ethnicity plays a crucial role in AD aetiology [50,51] and more diverse genomic datasets are needed to gain better unbiased insights [52].

In conclusion, we have established a ML approach for detecting genetic signals associated with disease, which goes some way to explain the missing heritability observed in previous literature.

## Methods

### Sample selection

Data for AD was obtained from two sources; ADNI and UKBB. The ADNI aimed at testing combinations of imaging and biological markers to measure progression of AD and mild cognitive impairment (MCI). For this study, cases were samples labelled as early and late MCI and AD (Supplementary Note 1). The UKBB contains phenotypic and biological information from 500,000 participants; see their previous publication for more details [21]. For this study, ICD10 codes from hospital inpatient records and participant responses were used to identify cases of AD. See supplementary for specific codes, question, and responses used. Additionally, individuals with indication of early onset AD and/or family history of AD were excluded. Based on the UK Biobank two subsets were generated to identify differences in detection power for novel variants. One contained a ratio of 1 case to 2 controls (labelled UKBB2to1) and the other a ratio of 1 case to 10 controls (labelled UKBB10to1). The UKBB10to1 cohort was used for all result sections, unless specified. Counts of individuals included in the analyses are shown in Supplementary Note 1. This research was approved by the UK Biobank’s governing Research Ethics Committee.

### Quality control

Quality control (QC) included exclusion of variants with minor allele frequency (MAF) < 0.01, imputation quality < 0.9, genotype missingness > 0.1 and those deviating from Hardy– Weinberg equilibrium (*P* < 1×10^-6^). Furthermore, individuals with a discrepancy between their genetic and reported sex were excluded and if their genotype-derived principal components 1 and 2 were further than 6 standard deviations away from those of 1000 Genomes European population. After QC, we had 11.7M variants in UKBB, and 9.5M variants in ADNI, with 4.6M in common between the two cohorts. Notably, the ADNI cohort was mapped to the GRCh38 reference while the UKBiobank was mapped to the GRCh37 reference.

### Genome-wide association study using logistic regression

Association testing between AD and genetic variants was conducted using whole genome LR model implemented in PLINK [53] (v1.90beta). Sex, age and the top 20 principal components were used as covariates for the association analysis.

### Genome-wide association study using VariantSpark

VariantSpark [19], a distributed implementation of the random forest (RF) algorithm, was used for association testing on Amazon Web Services. The same QC’d input files from LR analyses were used in the VariantSpark analyses. Optimisation of four hyperparameters; mTry, minNodeSize, MaxDepth, and nTree was run on all cohorts The optimised settings for all three cohorts were the same for mTry (0.1), MaxDepth (10), and nTree (20,000) except for minNodeSize where UKBB10to1 = 758, UKBB2to1 = 211, and ADNI = 78.

We determined the reliability of VariantSpark on real datasets by comparing Gini importance score of three runs on the UKBB10to1 and ADNI cohorts as Pearson’s correlations (Supplementary Figs. S1). Further, we tested the effect of covariates (as used in LR) in a RF model by comparing the out-of-bag error metric between a Ranger [54] run with covariates and a VariantSpark run without covariates. We did not observe any difference between the models; thus, covariates were not included in the final VariantSpark analysis.

### Compute resources

LR analyses were conducted on a machine with 16 Cores and 48 GiB memory. VariantSpark analyses were conducted using AWS Elastic Map Reduce with a total sum of 64 vCores and 488 GiB of memory.

### Post-GWAS analyses

#### P-value calculation

The primary measure of association from VariantSpark is the importance score derived from Gini-Index [55]. While this score can rank variants by importance, it is unable to determine significantly associated variants. To determine significance from importance scores, we used a recently developed method [22]. Briefly, this approach is based on the empirical Bayes method [56] which uses RF tree information as a threshold to fit a skew normal distribution and correct for multiple testing akin to Efron’s local false discovery rate approach.

#### Identification of independent variants, functional mapping and annotation

Variants identified in the GWAS were annotated using SNPTracker [57] and clumped using PLINK v.1.90b3.31 [53] within a window of 1,000kb and r^2^ of 0.01. Significantly associated variants were functionally mapped and annotated using ANNOVAR (v.7 2020-06-08) [58]. Furthermore, all significantly associated variants were mapped into locus bins where each locus bin was created based on a two million base-pair sliding window around the variants. This allowed known associations from the GWAS Catalog to be mapped to our results by identifying bins that are shared between the GWAS Catalog and our study’s associations.

#### General quality assurance of the UKBB (discovery) and ADNI (replication) cohort

PLINK LR results were used to identify potential population stratification using LDSC. No evidence for inflated statistics due to hidden population stratification was detected (LDSC intercept estimate was 1.03 ± 0.01 and 1.03 ± 0.01 for UKBB 10to1 and ADNI, respectively).

#### Epistasis calculation using BitEpi

To identify 2-SNP, 3-SNP, and 4-SNP interactions, BitEpi was applied to the significant VariantSpark associations in the UKBB and ADNI cohorts separately. The methods behind BitEpi have already been discussed elsewhere [59] but briefly, BitEpi calculates two entropy metrics, α and β. The β metric reflects the combined association power of all the SNPs involved in the interaction while the α metric represents the gain in association power due to the epistatic effect of all interactive SNPs. Therefore, an interaction with a large α and β has a strong association with the phenotype caused by an epistatic effect between all of the SNPs in the interaction. Quantiles for each order (2-SNP, 3-SNP or 4-SNP interactions) were used to filter out interactions with higher α and β values before *P*-values were computed through a permutation procedure. Bonferroni-corrected significance thresholds were calculated based on all possible combinations, with <0.05 denoting significance. SNPs involved in significant interactions were annotated with their independent SNP to remove any redundant interactions.

Using an in-house Python script, we generated contingency tables for some of the significant interactions found by BitEpi (Figure 4, Supplementary Table S9). The control rate is the number of controls over the number of samples for each genotype combination or for the overall cohort. The relative control rate is then the overall control rate minus the genotype combination control rate. A genotype combination with a negative relative control ratio can be considered to be deleterious and vice versa.

### Variance Explained Calculation

The significant associations from the VariantSpark, PLINK LR, and BitEpi analyses using the UKBB cohort were used to calculate the variance explained calculated as Nagelkerke’s pseudo-R2 [60] within the UKBB and ADNI cohort with the following as predictors in logistic models run using R v4.1.3 [61]; (1) significant and independent VariantSpark SNPs (*n* = 53), (2) significant and independent PLINK LR SNPs (*n* = 3), (3) significant and independent VariantSpark SNPs and all significant BitEpi interactions as interacting variables (*n* = 122). For all three models, the response was the AD case/control status. An empirical *P*-value was calculated from 1,000 ‘random noise’ models which were built to mimic the structure of model 3 by including the known *APOE* SNPs found by VariantSpark but also SNPs with no association with AD.

## Data Availability Statement

The data that support the findings of this study are available from ADNI database (https://adni.loni.usc.edu/data-samples/access-data/) and through the UK Biobank Data Showcase (http://www.ukbiobank.ac.uk/).

## Consortia for the Alzheimer’s Disease Neuroimaging Initiative

Michael W. Weiner^10^, Paul Aisen^11^, Ronald Petersen^12^, Clifford R. Jack^12^, Andrew J. Saykin^13^, William Jagust^14^, John Q. Trojanowki^15^, Arthur W. Toga^11^, Laurel Beckett^16^, Robert C. Green^17^, John Morris^18^, Leslie M. Shaw^15^, Zaven Khachaturian^19^, Greg Sorensen^20^, Maria Carrillo^21^, Lew Kuller^22^, Marc Raichle^18^, Steven Paul^23^, Peter Davies^24^, Howard Fillit^25^, Franz Hefti^26^, David Holtzman^18^, M. Marcel Mesulam^27^, William Potter^28^, Peter Snyder^29^, Adam Schwartz^30^, Tom Montine^31^, Ronald G. Thomas^32^, Michael Donohue^32^, Sarah Walter^32^, Devon Gessert^32^, Tamie Sather^32^, Gus Jiminez^32^, Archana B. Balasubramanian^32^, Jennifer Mason^32^, Iris Sim^32^, Danielle Harvey^16^, Matthew Bernstein^12^, Nick Fox^33^, Paul Thompson^34^, Norbert Schuff^10^, Charles Decarli^16^, Bret Borowski^12^, Jeff Gunter^12^, Matt Senjem^12^, David Jones^12^, Kejal Kantarci^12^, Chad Ward^12^, Robert A. Koeppe^35^, Norm Foster^36^, Eric M. Reiman^37^, Kewei Chen^37^, Chet Mathis^22^, Susan Landau^14^, Nigel J. Cairns^18^, Erin Franklin^18^, Lisa Taylor-Reinwald^18^, Virginia Lee^15^, Magdalena Korecka^15^, Michal Figurski^15^, Karen Crawford^11^, Scott Neu^11^, Tatiana M. Foroud^13^, Steven Potkin^38^, Kelley Faber^13^, Sungeun Kim^13^, Kwangsik Nho^13^, Leon Thal^32^, Neil Buckholtz^39^, Marilyn Albert^40^, Richard Frank^41^, John Hsiao^39^, Jeffrey Kaye^42^, Joseph Quinn^42^, Lisa Silbert^42^, Betty Lind^42^, Raina Carter^42^, Sara Dolen^42^, Lon S. Schneider^11^, Sonia Pawluczyk^11^, Mauricio Beccera^11^, Liberty Teodoro^11^, Bryan M. Spann^11^, James Brewer^32^, Helen Vanderswag^32^, Adam Fleisher^32^, Judith L. Heidebrink^35^, Joanne L. Lord^35^, Sara S. Mason^13^, Colleen S. Albers^12^, David Knopman^12^, Kris Johnson^12^, Rachelle S. Doody^43^, Javier Villanueva-Meyer^43^, Valory Pavlik^43^, Victoria Shibley^43^, Munir Chowdhury^43^, Susan Rountree^43^, Mimi Dang^43^, Yaakov Stern^44^, Lawrence S. Honig^44^, Karen L. Bell^44^, Beau Ances^18^, Maria Carroll^18^, Mary L. Creech^18^, Erin Franklin^18^, Mark A. Mintun^18^, Stacy Schneider^18^, Angela Oliver^18^, Daniel Marson^45^, David Geldmacher^45^, Marissa Natelson Love^45^, Randall Griffith^45^, David Clark^45^, John Brockington^45^, Erik Roberson^45^, Hillel Grossman^46^, Effie Mitsis^46^, Raj C. Shah^47^, Leyla Detoledo-Morrell^47^, Ranjan Duara^48^, Maria T. Greig-Custo^48^, Warren Barker^48^, Chiadi Onyike^40^, Daniel D’Agostino^40^, Stephanie Kielb^40^, Martin Sadowski^49^, Mohammed O. Sheikh^49^, Anaztasia Ulysse^49^, Mrunalini Gaikwad^49^, P. Murali Doraiswamy^50^, Jeffrey R. Petrella^50^, Salvador Borgesneto^50^, Terence Z. Wong^50^, Edward Coleman^50^, Steven E. Arnold^15^, Jason H. Karlawish^15^, David A. Wolk^15^, Christopher M. Clark^15^, Charles D. Smith^51^, Greg Jicha^51^, Peter Hardy^51^, Partha Sinha^51^, Elizabeth Oates^51^, Gary Conrad^51^, Oscar L. Lopez^22^, Mary Ann Oakley^22^, Donna M. Simpson^22^, Anton P. Porsteinsson^52^, Bonnie S. Goldstein^52^, Kim Martin^52^, Kelly M. Makino^52^, M. Saleem Ismail^52^, Connie Brand^52^, Adrian Preda^38^, Dana Nguyen^38^, Kyle Womack^53^, Dana Mathews^53^, Mary Quiceno^53^, Allan I. Levey^54^, James J. Lah^54^, Janet S. Cellar^54^, Jeffrey M. Burns^55^, Russell H. Swerdlow^55^, William M. Brooks^55^, Liana Apostolova^34^, Kathleen Tingus^34^, Ellen Woo^34^, Daniel H. S. Silverman^34^, Po H. Lu^34^, George Bartzokis^34^, Neill R Graff-Radford^56^, Francine Parfitt^56^, Kim Poki-Walker^56^, Martin R. Farlow^13^, Ann Marie Hake^13^, Brandy R. Matthews^13^, Jared R. Brosch^13^, Scott Herring^13^, Christopher H. Van Dyck^57^, Richard E. Carson^57^, Martha G. Macavoy^57^, Pradeep Varma^57^, Howard Chertkow^58^, Howard Bergman^58^, Chris Hosein^58^, Sandra Black^59^, Bojana Stefanovic^59^, Curtis Caldwell^59^, Ging-Yuek Robin Hsiung^60^, Benita Mudge^60^, Vesna Sossi^60^, Howard Feldman^60^, Michele Assaly^60^, Elizabeth Finger^61^, Stephen Pasternack^61^, Irina Rachisky^61^, John Rogers^61^, Dick Trost^61^, Andrew Kertesz^61^, Charles Bernick^62^, Donna Munic^62^, Emily Rogalski^27^, Kristine Lipowski^27^, Sandra Weintraub^27^, Borna Bonakdarpour^27^, Diana Kerwin^27^, Chuang-Kuo Wu^27^, Nancy Johnson^27^, Carl Sadowsky^63^, Teresa Villena^63^, Raymond Scott Turner^64^, Kathleen Johnson^64^, Brigid Reynolds^64^, Reisa A. Sperling^17^, Keith A. Johnson^17^, Gad Marshall^17^, Jerome Yesavage^65^, Joy L. Taylor^65^, Barton Lane^65^, Allyson Rosen^65^, Jared Tinklenberg^65^, Marwan N. Sabbagh^66^, Christine M. Belden^66^, Sandra A. Jacobson^66^, Sherye A. Sirrel^66^, Neil Kowall^67^, Ronald Killiany^67^, Andrew E. Budson^67^, Alexander Norbash^67^, Patricia Lynn Johnson^67^, Thomas O. Obisesan^68^, Saba Wolday^68^, Joanne Allard^68^, Alan Lerner^69^, Paula Ogrocki^69^, Curtis Tatsuoka^69^, Parianne Fatica^69^, Evan Fletcher^16^, Pauline Maillard^16^, John Olichney^16^, Charles Decarli^16^, Owen Carmichael^16^, Smita Kittur^70^, Michael Borrie^71^, T.-Y. Lee^71^, Rob Bartha^71^, Sterling Johnson^72^, Sanjay Asthana^72^, Cynthia M. Carlsson^72^, Pierre Tariot^37^, Anna Burke^37^, Ann Marie Milliken^37^, Nadira Trncic^37^, Adam Fleisher^37^, Stephanie Reeder^37^, Vernice Bates^73^, Horacio Capote^73^, Michelle Rainka^73^, Douglas W. Scharre^74^, Maria Kataki^74^, Brendan Kelly^74^, Earl A. Zimmerman^75^, Dzintra Celmins^75^, Alice D. Brown^75^, Godfrey D. Pearlson^76^, Karen Blank^76^, Karen Anderson^76^, Laura A. Flashman^77^, Marc Seltzer^77^, Mary L. Hynes^77^, Robert B. Santulli^77^, Kaycee M. Sink^78^, Leslie Gordineer^78^, Jeff D. Williamson^78^, Pradeep Garg^78^, Franklin Watkins^78^, Brian R. Ott^79^, Geoffrey Tremont^79^, Lori A. Daiello^79^, Stephen Salloway^80^, Paul Malloy^80^, Stephen Correia^80^, Howard J. Rosen^10^, Bruce L. Miller^10^, David Perry^10^, Jacobo Mintzer^81^, Kenneth Spicer^81^, David Bachman^81^, Nunzio Pomara^82^, Raymundo Hernando^82^, Antero Sarrael^82^, Susan K. Schultz^83^, Karen Ekstam Smith^83^, Hristina Koleva^83^, Ki Won Nam^83^, Hyungsub Shim^83^, Norman Relkin^23^, Gloria Chaing^23^, Michael Lin^23^, Lisa Ravdin^23^, Amanda Smith^84^, Balebail Ashok Raj^84^ & Kristin Fargher^84^

^10^University Of California, San Francisco, USA. ^11^University Of Southern California, Los Angeles, USA. ^12^Mayo Clinic, Rocester, USA ^13^Indiana University, Bloomington, USA. ^14^University Of California, Berkeley, Berkeley, USA. ^15^University Of Pennsylvania, Philadelphia, USA. ^16^University Of California, Davis, Davis, USA. ^17^Brigham And Women’s Hospital/Harvard Medical School, Boston, USA. ^18^Washington University St. Louis, St. Louis, USA. ^19^Prevent Alzheimer’s Disease, 2020, Rockville, USA. ^20^Siemens, Munich, Germany. ^21^Alzheimer’s Association, Illinois, USA. ^22^University Of Pittsburgh, Pennsylvania, USA. ^23^Cornell University, New York, USA. ^24^Albert Einstein College of Medicine of Yeshiva University, New York, USA. ^25^AD Drug Discovery Foundation, New York, USA. ^26^Acumen Pharmaceuticals, California, USA. ^27^Northwestern University, Illinois, USA. ^28^National Institute of Mental Health, Maryland, USA. ^29^Brown University, Rhode Island, USA. ^30^Eli Lilly, Indiana, USA. ^31^University Of Washington, Washington, USA. ^32^University Of California, San Diego, California, USA. ^33^University Of London, London, UK. ^34^University Of California, Los Angeles, California, USA. ^35^University Of Michigan, Michigan, USA. ^36^University Of Utah, Utah, USA. ^37^Banner Alzheimer’s Institute, Arizona, USA. ^38^University Of California, Irvine, California, USA. ^39^National Institute on Aging, Maryland, USA. ^40^Johns Hopkins University, Maryland, USA. ^41^Richard Frank Consulting, New Hampshire, USA. ^42^Oregon Health and Science University, Oregon, USA. ^43^Baylor College Of Medicine, Texas, USA. ^44^Columbia University Medical Center, New York, USA. ^45^University Of Alabama-Birmingham, Alabama, USA. ^46^Mount Sinai School of Medicine, New York, USA. ^47^Rush University Medical Center, Rush University, Illinois, USA. ^48^Wien Center, Florida, USA. ^49^New York University, New York, USA. ^50^Duke University Medical Center, North Carolina, USA. ^51^University Of Kentucky, Kentucky, USA. 10University Of California, San^52^University Of Rochester Medical Center, New York, USA. ^53^University Of Texas Southwestern Medical School, Texas, USA. ^54^Emory University, Georgia, USA. ^55^University Of Kansas, Medical Center, Kansas, USA. ^56^Mayo Clinic, Jacksonville, Florida, USA. ^57^Yale University School of Medicine, Connecticut, USA. ^58^Mcgill University, Montreal-Jewish General Hospital, Quebec, Canada. ^59^Sunnybrook Health Sciences, Ontario, Canada. ^60^U.B.C. Clinic for AD & Related Disorders, British Columbia, Canada. ^61^Cognitive Neurology-St. Joseph’s, Ontario, Canada. ^62^Cleveland Clinic Lou Ruvo Center for Brain Health, Ohio, USA. ^63^Premiere Research Inst (Palm Beach Neurology), Florida, USA. ^64^Georgetown University Medical Center, Washington, D.C, USA. ^65^Stanford University, California, USA. ^66^Banner Sun Health Research Institute, Arizona, USA. ^67^Boston University, Massachusetts, USA. ^68^Howard University, Washington, D.C, USA. ^69^case Western Reserve University, Ohio, USA. ^70^Neurological Care Of CNY, New York, USA. ^71^Parkwood Hospital, Pennsylvania, USA. ^72^University Of Wisconsin, Wisconsin, USA. ^73^Dent Neurologic Institute, New York, USA. ^74^Ohio State University, Ohio, USA. ^75^Albany Medical College, New York, USA. ^76^Hartford Hospital, Olin Neuropsychiatry Research Center, Connecticut, USA. ^77^Dartmouth-Hitchcock Medical Center, New Hampshire, USA. ^78^Wake Forest University Health Sciences, North Carolina, USA. ^79^Rhode Island Hospital, Rhode Island, USA. ^80^Butler Hospital, Rhode Island, USA. ^81^Medical University South Carolina, Carolina, USA. ^82^Nathan Kline Institute, New York, USA. ^83^University Of Iowa College of Medicine, Iowa, USA. ^84^USF Health Byrd Alzheimer’s Institute, University of South Florida, Florida, USA.

## Acknowledgements

Funding was from CSIRO Health and Biosecurity. The results published here are in whole or in part based on data obtained from Agora, a platform initially developed by the NIA-funded AMP-AD consortium that shares evidence in support of AD target discovery. Data collection and sharing for this project was funded by the Alzheimer’s Disease Neuroimaging Initiative (ADNI) (National Institutes of Health Grant U01 AG024904) and DOD ADNI (Department of Defense award number W81XWH-12-2-0012). ADNI is funded by the National Institute on Aging, the National Institute of Biomedical Imaging and Bioengineering, and through generous contributions from the following: AbbVie, Alzheimer’s Association; Alzheimer’s Drug Discovery Foundation; Araclon Biotech; BioClinica, Inc.; Biogen; Bristol-Myers Squibb Company; CereSpir, Inc.; Cogstate; Eisai Inc.; Elan Pharmaceuticals, Inc.; Eli Lilly and Company; EuroImmun; F. Hoffmann-La Roche Ltd and its affiliated company Genentech, Inc.; Fujirebio; GE Healthcare; IXICO Ltd.; Janssen Alzheimer Immunotherapy Research & Development, LLC.; Johnson & Johnson Pharmaceutical Research & Development LLC.; Lumosity; Lundbeck; Merck & Co., Inc.; Meso Scale Diagnostics, LLC.; NeuroRx Research; Neurotrack Technologies; Novartis Pharmaceuticals Corporation; Pfizer Inc.; Piramal Imaging; Servier; Takeda Pharmaceutical Company; and Transition Therapeutics. The Canadian Institutes of Health Research is providing funds to support ADNI clinical sites in Canada. Private sector contributions are facilitated by the Foundation for the National Institutes of Health (www.fnih.org). The grantee organization is the Northern California Institute for Research and Education, and the study is coordinated by the Alzheimer’s Therapeutic Research Institute at the University of Southern California. ADNI data are disseminated by the Laboratory for Neuro Imaging at the University of Southern California. Samantha Burnham is now employed at “Avid Radiopharmaceuticals, a wholly owned subsidiary of Eli Lilly and Company.”

## Author contributions

N.A.T, D.C.B, M.L conceptualised the study. L.M.F.S, P.S, R.D, M.L, A.B, N.A.T, D.C.B contributed to primary data analysis, including statistical analysis. N.A.T, D.C.B, M.L, L.M.F.S wrote the manuscript. S.C.B contributed to the interpretation of the data. All authors critically reviewed and gave final approval for the manuscript.

