## Supplementary Notes and Figures for "Novel Alzheimer’s disease genes and epistasis identified using machine learning GWAS platform"

**Supplementary Note 1**

UK Biobank (UKBB) Samples

The hospital inpatient records and participant responses connected to the UKBB were used to select cases and controls for this study. Specifically, the following ICD10 codes were used to determine cases of Alzheimer’s disease (AD):

- F00.1: Dementia in Alzheimer’s disease with late onset
- F00.2 Dementia in Alzheimer’s disease, atypical or mixed type
- F00.9: Dementia in Alzheimer’s disease, unspecified
- G30.1: Alzheimer’s disease with late onset
- G30.8: Other Alzheimer’s disease
- G30.9: Alzheimer’s disease, unspecified

Furthermore, participant responses from the following questions were also used:

- Illnesses of the father
- Illnesses of the mother
- Illnesses of the sibling

**Supplementary Note 1, Table 1.** Cohort details with the number of cases and controls used in this study. ADNI refers to the Alzheimer’s disease neuroimaging initiative and UKBB to the UK Biobank. EMCI refers to early mild cognitive impairment and LMCI to late mild cognitive impairment.

| Cohort | Phenotype | Cases | | | Controls | | | All | | |
| --- | --- | --- | --- | --- | --- | --- | --- | --- | --- | --- |
|  |  | Male | Female | Total | Male | Female | Total | Male | Female | Total |
| ADNI | AD, EMCI, LMCI | 305 | 207 | 512 | 135 | 137 | 272 | 440 | 344 | 784 |
| UKBB2to1 | AD | 370 | 334 | 704 | 739 | 667 | 1406 | 1109 | 1001 | 2110 |
| UKBB10to1 | AD | 370 | 334 | 704 | 3611 | 3258 | 6869 | 3981 | 3592 | 7573 |

**Supplementary Note 1, Table 2.** Raw counts of ADNI participants used as AD cases and/or excluded. Note that the participants between the ADNI AD types might overlap and therefore should not be summed. AD: Alzheimer’s disease. EMCI: Early mild cognitive impairment. LMCI: Late mild cognitive impairment. SMC: Significant memory concern.

| ADNI AD Type | AD | EMCI | LMCI | SMC |
| --- | --- | --- | --- | --- |
| Cases | 45 | 230 | 237 | 0 |

**Supplementary Note 1, Table 3.** Raw counts of UKBB participants used as AD cases and/or excluded. Note that the participants between the different UKBB data field IDs might overlap and therefore should not be summed. Codes in data fields 41202 and 41204 are explained in the supplementary methods.

| UKBB Data Field ID | Included/Excluded | Codes | Number of Participants |
| --- | --- | --- | --- |
| 20107: Illnesses of Father | Excluded | NA | 26,609 |
| 20110: Illnesses of Mother | Excluded | NA | 50,083 |
| 20111: Illnesses of Sibling | Excluded | NA | 3,241 |
| 41202: ICD10 (Primary) | Excluded | F00.0 | 16 |
|  | Included | F00.1 | 21 |
|  | Included | F00.2 | 31 |
|  | Included | F00.9 | 50 |
|  | Excluded | G30.0 | 38 |
|  | Included | G30.0 | 43 |
|  | Included | G30.2 | 93 |
|  | Included | G30.9 | 257 |
| 41204: ICD10 (Secondary) | Excluded | F00.0 | 102 |
|  | Included | F00.1 | 69 |
|  | Included | F00.2 | 375 |
|  | Included | F00.9 | 1,582 |
|  | Excluded | G30.0 | 157 |
|  | Included | G30.0 | 57 |
|  | Included | G30.2 | 337 |
|  | Included | G30.9 | 2,185 |

**Supplementary Figures**

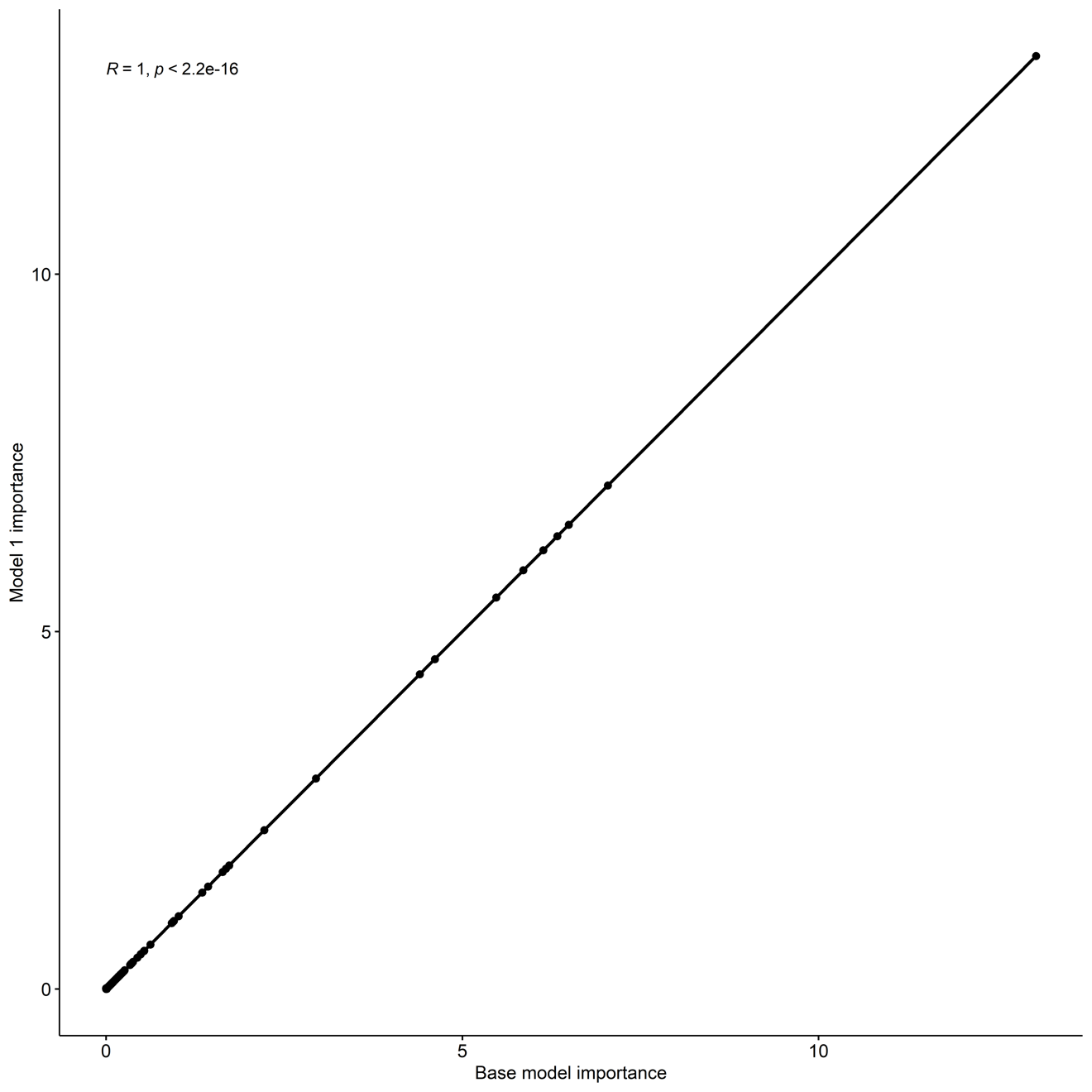

**Supplementary Figure 1A.** Pearson’s correlation plot comparing the Gini importance scores for 5,337,197 SNPs for 2 separate VariantSpark models with the same hyperparameters but different random seeds. This was performed using the UK Biobank cohort (10 controls to 1 case). In total 3 different models were generated, each with different random seeds. This plot compares the base model and model 1.

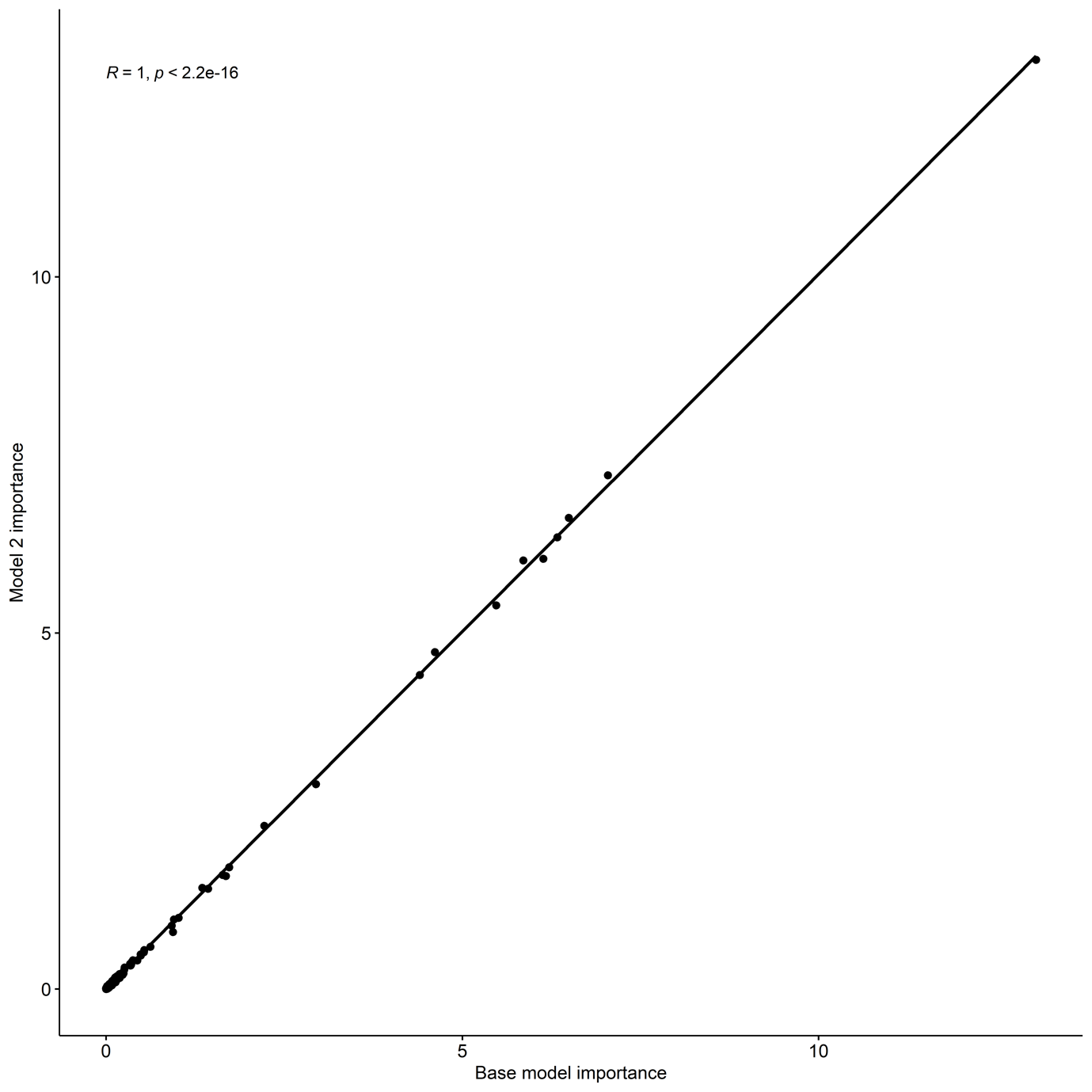

**Supplementary Figure 1B.** Pearson’s correlation plot comparing the Gini importance scores for 5,337,197 SNPs for 2 separate VariantSpark models with the same hyperparameters but different random seeds. This was performed using the UK Biobank cohort (10 controls to 1 case). In total 3 different models were generated, each with different random seeds. This plot compares the base model and model 2.

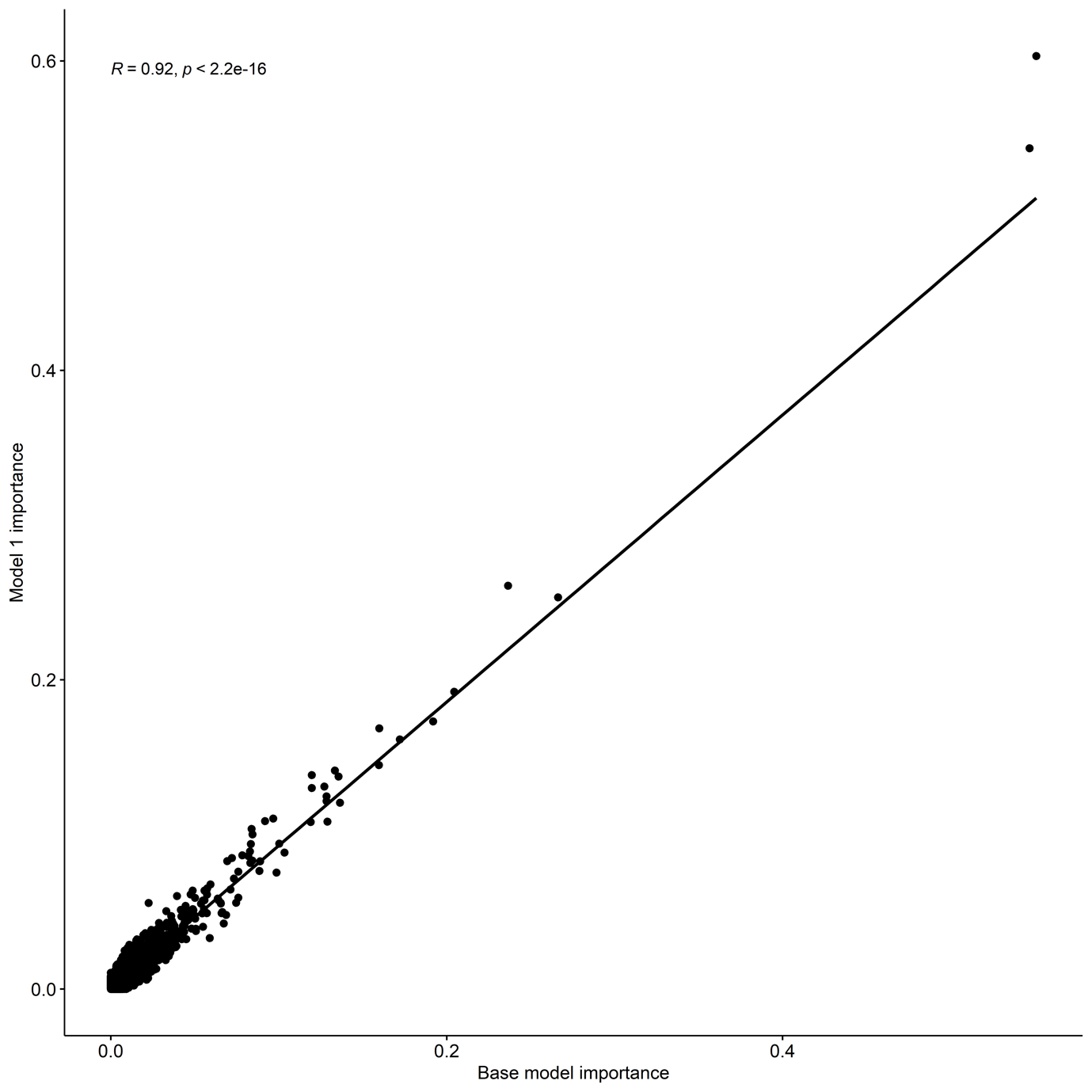

**Supplementary Figure 1C.** Pearson’s correlation plot comparing the Gini importance scores for 4,552,760 SNPs for 2 separate VariantSpark models with the same hyperparameters but different random seeds. This was performed using the ADNI cohort. In total 3 different models were generated, each with different random seeds. This plot compares the base model and model 1.

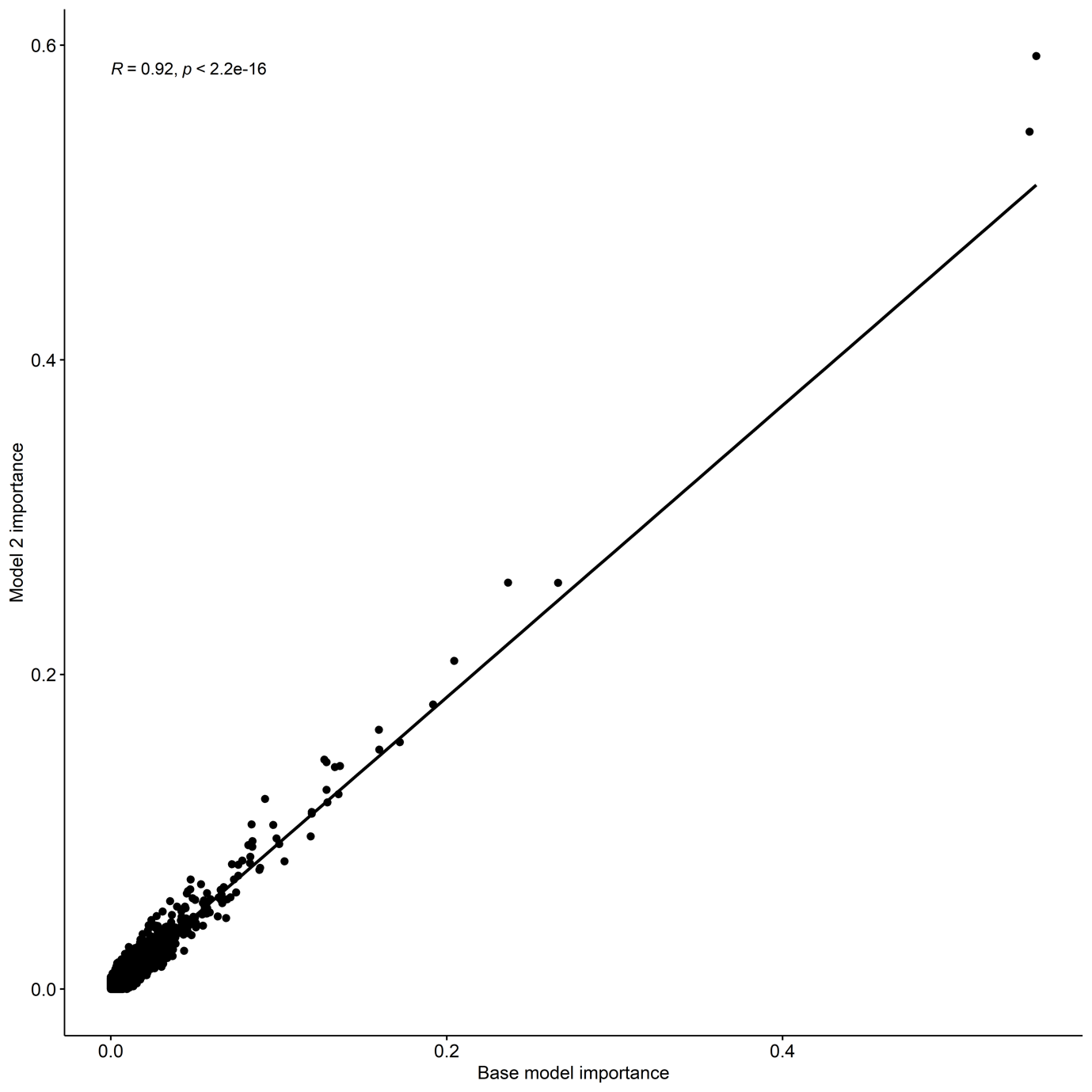

**Supplementary Figure 1D.** Pearson’s correlation plot comparing the Gini importance scores for 4,552,760 SNPs for 2 separate VariantSpark models with the same hyperparameters but different random seeds. This was performed using the ADNI cohort. In total 3 different models were generated, each with different random seeds. This plot compares the base model and model 2. 

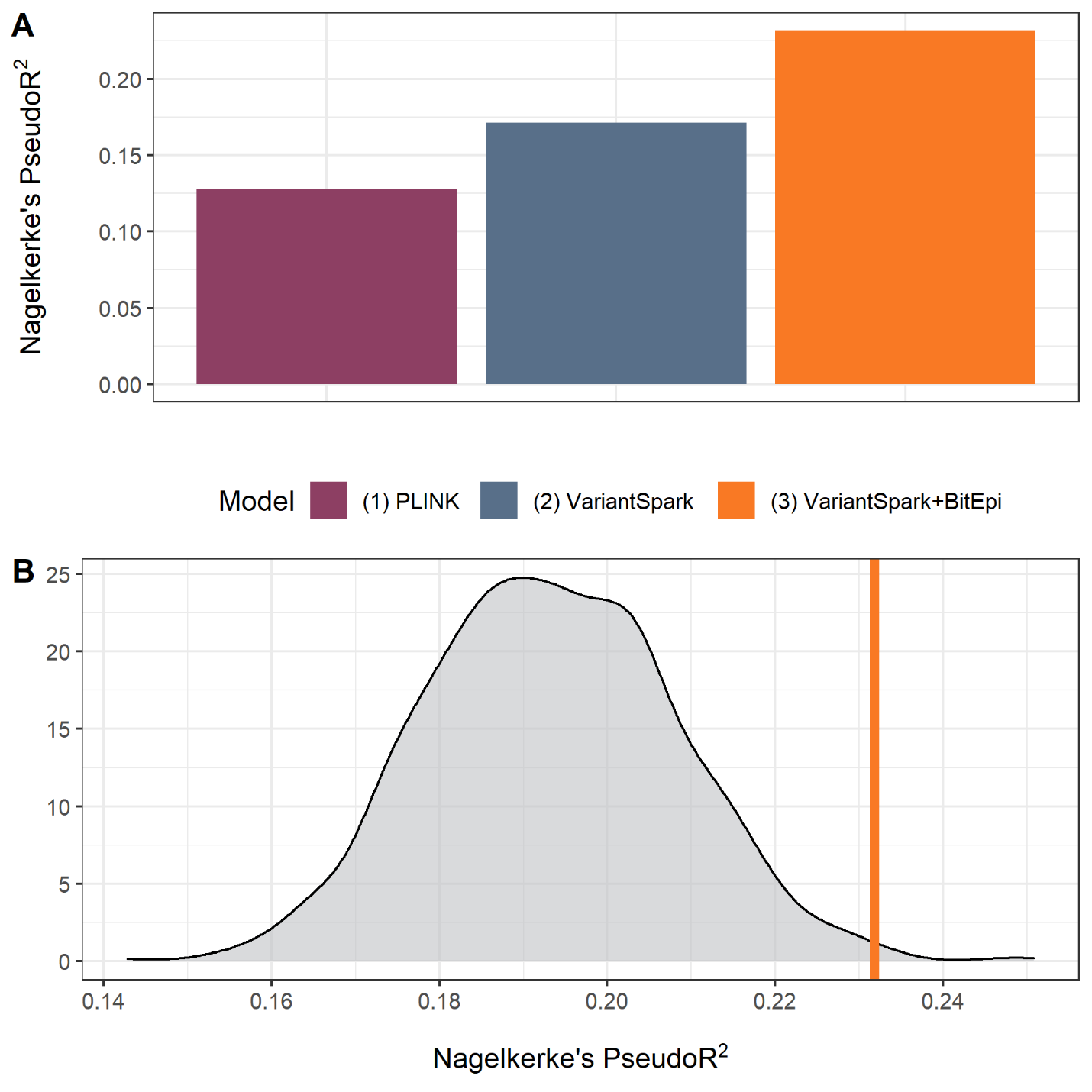

**Supplementary Figure 2**. (A) Variance explained calculated as Nagelkerke’s PseudoR^2^ for three models: (1) PLINK Logistic Regression SNPs, (2) VariantSpark SNPs, (3) VariantSpark SNPs + BitEpi Interactions. The model with VariantSpark SNPs + BitEpi interactions had the largest R^2^, followed by the model with VariantSpark SNPs only, and lastly by the model with PLINK SNPs. This indicates that epistasis can explain more phenotypic variance than marginal associations alone. (B) Density plot of Nagelkerke’s PseudoR^2^ of 1,000 models built with same model structure as model (3) VariantSpark SNPs + BitEpi interactions but using random non-associated SNPs. Vertical line indicates the R^2^ of model (3) = 0.23 with a calculated empirical *P* value = 0.006.

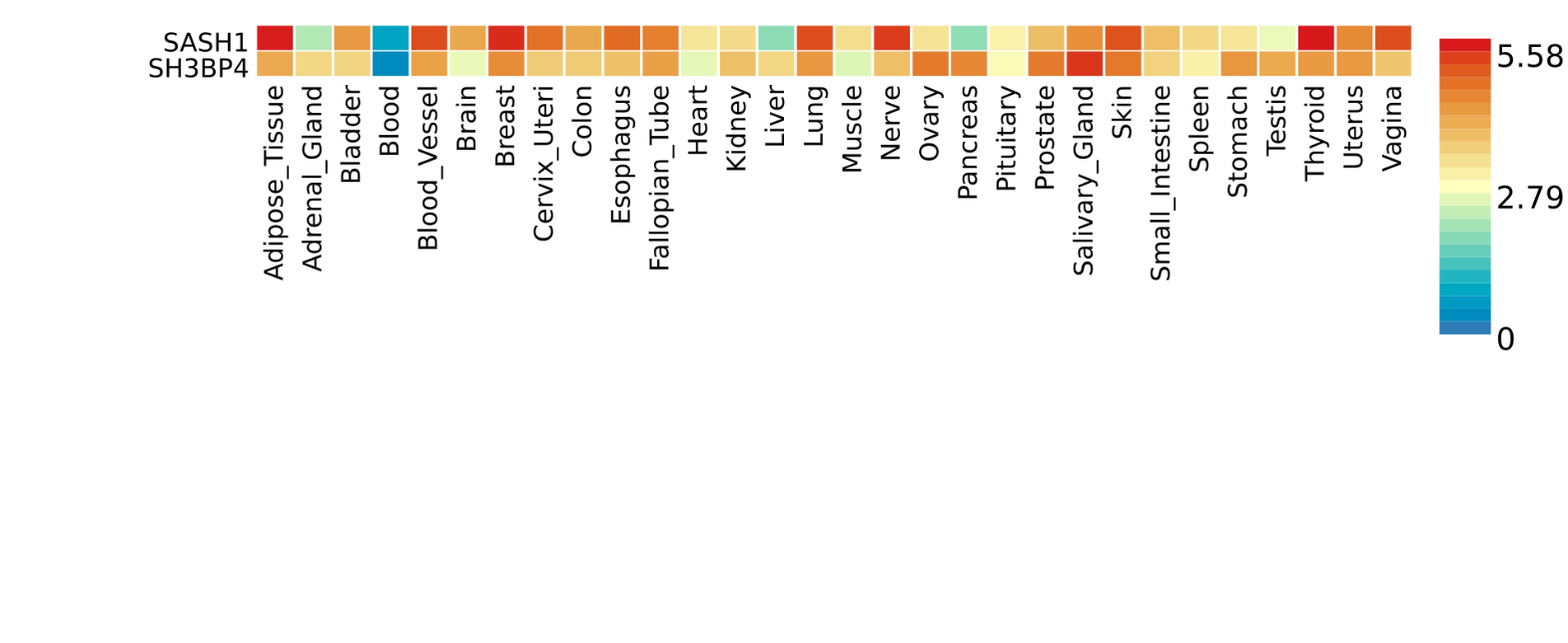

**Supplementary Figure 3**. Gene expression heatmap output from FUMA’s GENE2FUNC for *SASH1* and *SH3BP4*. The cells of the heatmap represent the averaged expression value per tissue type for each gene following winsorization at 50 and log 2 transformation with pseudocount 1 where cells coloured red represent higher expression compared to cells filled in blue across genes and tissue type. Both *SASH1* and *SH3BP4* have strong expression in brain tissue.
